# Validation of a Difference-in-Differences Investigation Tool (DiD IT) for quantifying local outbreaks

**DOI:** 10.1101/2024.09.20.24314075

**Authors:** Roger Morbey, Andre Charlett, Daniel Todkill, Alex J. Elliot

## Abstract

The Difference-in-Differences Investigation Tool (‘DiD IT’) is a new tool used to estimate the impact of local threats to public health in England. ‘DiD IT’ is part of a daily all hazards syndromic surveillance service. We present a validation of the ‘DiD IT’ tool, using synthetic injects to assess how well it can estimate small, localised increases in the number of people presenting to health care. Furthermore, we assess how control settings within ‘DiD IT’ affect it’s performance.

‘DiD IT’ was validated across ten different syndromic indicators, chosen to cover a range of data volumes and potential public health threats. Injects were added across different times of year and days of week, including public holidays. Also, different size of injects were created, including some with an impact spread to neighbouring locations or spread over several days. The control settings within ‘DiD IT’ were tested by varying the control location and periods, using, for example a ‘washout period’ or excluding nearest neighbours. Performance was measured by comparing the estimates for excess counts produced by ‘DiD IT’ with the actual synthetic injects added.

‘DiD IT’ was able to provide a positive estimate in 99.8% of trials, with a mean absolute error of 1.5. However, confidence intervals for the central estimate could not be produced in 42.5% of trials. Furthermore, the 95% confidence intervals for the central estimates only included the actual inject count within 62.8% of the intervals. Unsurprising, mean errors were slightly higher when synthetic injects were not concentrated in one location on one day but were spread across neighbouring areas or days. Selecting longer control periods and using more locations as controls tended to lower the errors slightly. Including a washout period or excluding neighbouring locations from the controls did not improve performance.

We have shown that ‘DiD IT’ is accurate for assessing the impact of local incidents but that further work is needed to improve the how the uncertainty of these estimates are communicated to users.

## Introduction

The Difference-in-Differences Investigation Tool (‘DiD IT’) was developed utilising real-time syndromic surveillance data to support risk assessments of the impact of local threats to public health [1]. ‘DiD IT’ estimates the excess number of cases reported daily to syndromic surveillance systems, due to a localised outbreak or incident.

‘DiD IT’ uses a difference-in-differences statistical approach to account for temporal and spatial confounding and provide a direct estimate of impact due to incidents.

Temporal confounding differences are estimated by comparing unaffected locations during and outside of exposure periods. Whilst spatial confounding differences are estimated by comparing unaffected and exposed locations outside of the exposure period. Therefore, any remaining differences after accounting for temporal and spatial confounding are the direct effect of the local incident.

Importantly, ‘DiD IT’ is not an outbreak detection tool, but is designed to support situational awareness (one of the key aims of syndromic surveillance [2]); estimating the size of impact when the potential exposure period and location are already known by statistically comparing with similar time periods and geographies related to exposures with those thought not to impacted. To further establish the usefulness of this tool we need to validate the accuracy of ‘DiD IT’ so that incident directors and decision makers can have confidence in its use for situational awareness.

The UK Health Security Agency (UKHSA) real-time syndromic surveillance programme includes the daily monitoring of over 130 different syndromic indicators across six national syndromic surveillance systems [3]. The indicators have been developed to monitor as wide a range as possible of potential public threats i.e. an ‘all hazards approach [4]. Thus, ‘DiD IT’ can be applied to syndromic data in scenarios ranging from environmental incidents to mass gatherings or infectious disease outbreaks. However, many potential public health scenarios are rare and therefore little or no relevant historical data are available. Therefore, in this study we have validated ‘DiD IT’ using synthetic injects, adding simulated events to real-time series of syndromic daily counts. Using simulated data enables us to test a wide range of potential scenarios, and identify factors, such as public holidays, that might affect accuracy of the tool. We also test how different options for selecting controls affect the sensitivity of estimates created by ‘DiD IT’. Thus, we aim to provide future guidelines for setting controls that maximise accuracy.

## Methods

### Data extraction

The UKHSA real-time syndromic surveillance programme coordinates and monitors daily data from six national (England) systems that has been previous described [5-7]. Here, we extracted daily counts for 10 syndromic indicators across five syndromic systems to represent the range of data used for daily surveillance (**Table 1**). For each system we chose two syndromic indicators, one with more counts on average than the other.

**Table 1:** Syndromic surveillance systems and syndromic indicators selected for use in the study.

| Syndromic data source | Syndromic indicator |
| --- | --- |
| General practitioner in-hours consultations | Gastroenteritis |
|  | Vomiting |
| General practitioner out-of-hours consultations | Eye problems |
|  | Respiratory |
| Ambulance dispatch calls | Allergic reactions |
|  | Breathing problems |
| National Health Service (NHS) 111 telehealth calls | Eye problems |
|  | Cough |
| National Health Service (NHS) 111 online symptom checker | Eye problems |
|  | Cough |

Counts for each syndromic indicator were aggregated to postcode district area level (which was the lowest geographical area available). Data were extracted for the period April 2021 to August 2022 inclusive, which was when data was available for all five systems. The general practitioner (GP) surveillance systems (in-hours and out-of-hours) are sentinel providing partial population coverage whilst the other systems cover the whole population of England. Dates were randomly selected to be used as the centre of exposure periods. One date was purposely randomly selected from the public holidays between April 2021 and August 2022. The other dates were randomly selected from dates that were not on public holidays and far enough away from public holidays that both exposure and control periods would not include public holidays. Twenty-one non-holiday dates were used, sampled as three random days plus the six preceding dates for each of the three. Thus, we were able to compare the accuracy of ‘DiD IT’ by day of the week and measure any impact of public holidays. Exposure locations were selected randomly from postcode districts that had at least one non-zero count for the syndromic indicator being tested during the exposure and control periods.

### Synthetic injects

We created synthetic injects to simulate local incidents. To simulate different types of incidents, a range of injects were created. Two sizes of inject were created, a ‘small’ inject with a peak count of five cases, and ‘large’ with a peak of 15 cases. Also, injects were created to simulate different temporal and geographical spreads. Half of the injects were treated as single day exposures, the other half were spread symmetrically over five consecutive days, with the peak in cases on the middle date. Similarly, half the injects involved a single postcode district, whilst the other half included a peak in one district with lower numbers in surrounding districts. The combination of two sizes, two temporal and two geographical spreads gave eight different inject types.

For each of the twenty-two dates randomly selected as described above a time series was created for each syndromic indicator using actual daily counts. For each of these series, eight new time series were created, each with additional counts from one of the eight different sizes and spreads of synthetic injects.

### Control options

A range of different rules for selecting controls were tested. Four different options for control period were tested, either seven days long or the same as the exposure period (1 or 5 days depending on inject spread), and with or without a ‘washout’ period of seven days between control and exposure periods. Similarly, three different options were tested for selecting control locations; using all available non-exposed postcode districts, using all except those bordering the exposure locations and using ten random districts that did not border the exposure.

For the injects that were spread over several days, two exposure periods were tested, firstly the full five days and second just using the middle peak day. Similarly for injects that included neighbouring districts, two exposure locations were considered, just the central district or the district and its neighbours. Including these alternate options for exposure simulated incidents where the central location and date were known but the spread had not been identified.

The combination of different exposure dates, syndromic indicators, inject types and control options resulted in 60,672 separate ‘trials’ used to validate ‘DiD IT’.

### DiD IT

We applied ‘DiD IT’ to each of the separate 60,672 trials and recorded the number of extra cases estimated along with a confidence interval for the parameter estimate.

The exposure location(s) and period were determined by the inject type, whilst control period and locations determined by the trial’s control options. ‘DiD IT’ was applied using a negative binomial regression to predict the daily syndromic count for each postcode district. Three binary variables were created as the independent variables; exposure period, exposure location and effect incident, ‘exposure period’ was a 1 for dates during the exposure period, ‘exposure location’ was a 1 for the exposed districts and ‘effect incident’ was the product of the first two variables. Thus, ‘effect incident’ was a parameter used to estimate the additional cases due to an incident. For each trial the error was defined as the difference between the estimate for ‘effect incident’ and the number of extra cases added by the synthetic inject.

## Results

The regression method used by ‘DiD IT’ does not always converge, due to sparse data, in which case no estimate can be provided. Also, on rare occasions the estimate for number of extra cases due to an incident may be negative, which can be considered as a failure to provide a positive estimate. However, 60,540 of the trials (99.8%) resulted in a positive estimate for the number of excess cases.

Importantly, ‘DiD IT’ does not just provide an estimate for the number of extra cases but tries to illustrate the uncertainty around this estimate by also providing a confidence interval. The current method for calculating confidence intervals did not converge for 25,766 trials, (42.5% of the total). For 10,406 trials (17.2%) a 95% confidence interval was calculated but it spanned zero. For the 24,399 trials (40.3%) where the lower confidence interval was positive, 15,323 (62.8% of those with a CI) correctly included the inject total within the 95% interval. For these confidence intervals that did not span zero, the mean interval length was 11.57, with half of the intervals being 7.77 or less. The widest confidence interval was where the estimate was 124.4 excess counts, 95% confidence interval (11.3 – 179.5), in this trial the actual inject total was 106 spreading over 5 days, including neighbouring districts. Thirty-three trials resulted in confidence intervals where the upper estimate was over 100 more than the positive lower estimate.

The average absolute error in estimates was 1.50, with an interquartile range for the errors of -0.64 to 0.58. The underestimate that was least accurate was 129.8 (95% CI 77.3 – 162.2) where 171 extra counts were injected, and the least accurate overestimate was 183 (95% CI 158.4 – 195.0) when 145 extra had been injected.

### Syndromic indicator factors

When considering daily data at the smallest geographical area available the mean count is low, with many zeroes. For each system the mean absolute error was greater for the syndrome with higher counts. For the rarer syndromic indicators, the errors were less than 1 for each system (**Table 2**). For the more common syndromic indicators, mean absolute errors ranged from 1.78 for NHS 111 online cough assessments to 2.66 for NHS 111 cough calls. The GP in-hours system resulted in less trials where a positive estimate could be obtained. Unlike the other systems, GP in-hours is a five-day system with nearly no data available for weekends and public holidays.

**Table 2:** Mean absolute error by syndromic indicator.

| Syndromic data source | Syndromic indicator | Daily count in postcode district |  | Number of trials with positive estimates | Mean error (absolute difference between estimate and extra cases injected) |
| --- | --- | --- | --- | --- | --- |
|  |  | Mean | Max |  |  |
| GP in-hours | Gastroenteritis | 0.79 | 20 | 4,966 | 2.50 |
|  | Vomiting | 0.13 | 7 | 4,985 | 0.65 |
| GP out-of-hours | Eye problems | 0.08 | 4 | 6,336 | 0.30 |
|  | Respiratory conditions | 1.15 | 48 | 6,296 | 2.57 |
| Ambulance dispatch calls | Allergic reactions | 0.10 | 5 | 6,326 | 0.50 |
|  | Breathing problems | 0.96 | 36 | 6,318 | 2.29 |
| NHS 111 calls | Eye problems | 0.31 | 11 | 6,334 | 0.79 |
|  | Cough | 0.94 | 24 | 6,317 | 2.66 |
| NHS 111 online | Eye problems | 0.17 | 10 | 6,335 | 0.98 |
|  | cough | 0.44 | 12 | 6,327 | 1.78 |
| **All syndromic indicators** |  |  |  | 60,540 | 1.50 |

### Inject factors

Unlike the size of the daily counts in each syndromic indicator, the size of the inject had little impact on the mean absolute error, small injects had an average error of 1.48, large 1.51 (**Table 3**).

**Table 3:** Absolute mean error by inject factor.

| **Inject factor** | **Number of trials with positive estimates** | **Average error (absolute difference between estimate and extra cases injected)** |
| --- | --- | --- |
| **Size of inject – number of cases at peak** |  |  |
| 5 | 30,215 | 1.48 |
| 15 | 30,325 | 1.51 |
| **Temporal spread of inject** |  |  |
| Single day | 19,732 | 0.93 |
| Five days, test peak day only | 19,718 | 0.93 |
| Five days exposure period | 21,090 | 2.55 |
| **Geographical spread of inject** |  |  |
| Single district | 20,172 | 0.99 |
| District and neighbours, test central district only | 20,171 | 0.99 |
| District and neighbours used for exposure | 20,197 | 2.51 |
| **Day of the week of peak exposure** |  |  |
| Public holiday | 2,490 | 1.83 |
| Saturday | 7,465 | 1.63 |
| Sunday | 7,484 | 1.56 |
| Monday | 8,617 | 1.49 |
| Tuesday | 8,623 | 1.48 |
| Wednesday | 8,596 | 1.49 |
| Thursday | 8,633 | 1.35 |
| Friday | 8,632 | 1.39 |

Errors were higher when the exposure was not focussed on a single day or location. The magnitude of the increase in errors was very similar for geography and temporal spread. The mean absolute error for trials where the exposure was a single day was 0.93 and 0.99 for a single postcode district, rising to 2.55 for a five-day exposure and 2.51 for an exposure including neighbouring districts. These results were cumulative, so that the error for trials involving a single exposure date and district was 0.57 and for trials with an exposure period of five days and neighbours being 4.12. The accuracy of peak estimates for the central date or central district were very similar, whether the inject involved temporal or geographical spread.

The accuracy of ‘DiD IT’ was slightly worse when the inject was on a public holiday, mean absolute error of 1.83 and weekends had slightly higher errors then other days, with Thursdays and Fridays having the lowest errors of 1.35 and 1.39 (**Table 3**).

### Control factors

The mean absolute errors were slightly less when a full seven days were used for the control period, as opposed to being the same as the exposure period (1 or 5 days). Also, errors were slightly lower when the control period was immediately preceding the exposure period, rather than with a ‘washout’ period of seven days between control and exposure (**Table 4**).

**Table 4:** Absolute mean error by control factor.

| Control factor | Number of trials with positive estimates | Average error (absolute difference between estimate and extra cases injected) |
| --- | --- | --- |
| **Control period** |  |  |
| 7 days before exposure | 15,159 | 1.32 |
| 7 days with washout of 7 days before exposure | 15,166 | 1.49 |
| Same length as exposure | 15,116 | 1.55 |
| Same length as exposure with washout of 7 days | 15,099 | 1.63 |
| **Control locations** |  |  |
| Non-neighbouring districts | 15,146 | 1.38 |
| All non-exposed districts | 15,152 | 1.39 |
| Ten random non-neighbours | 30,242 | 1.61 |

Mean absolute errors were higher (1.61) when only 10 districts were used as controls. Excluding neighbouring districts made no significant difference when using all available control districts.

The biggest variation in errors was between syndromic indicators, followed by inject factors. By contrast the differences between control factors or day of the week were less important (**Figure 1**).

**Figure 1:**
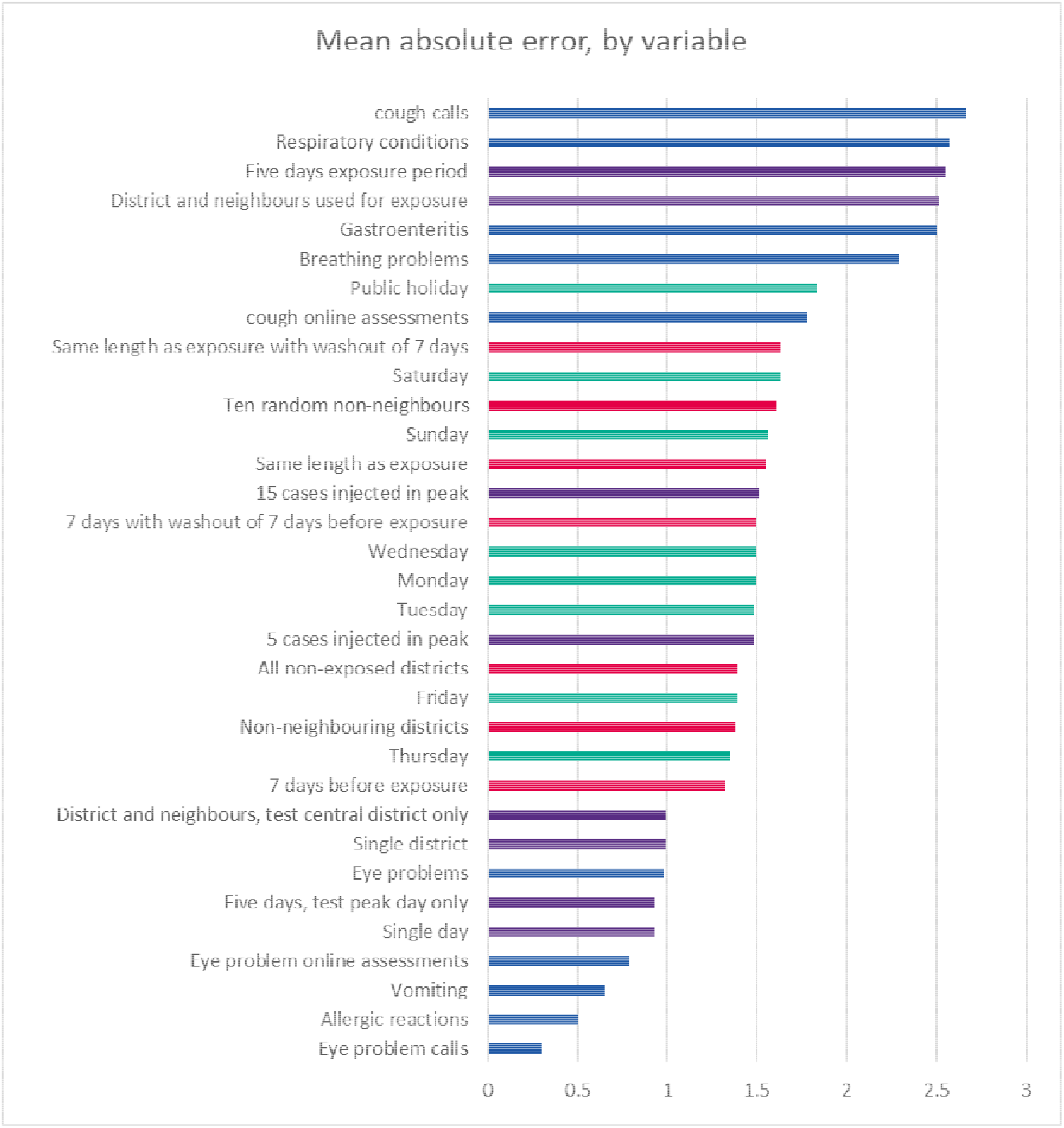
syndromic indicators shown in blue, inject factors in purple, control factors in red and day of week in teal.

## Discussion

### Key findings

‘DiD IT’ was able to estimate the number of extra cases due to the synthetic incident with a high level of accuracy, the mean error was 1.5 with most estimates being less than 1 away from the total added. However, the current method for calculating uncertainty around the estimates needs improvement. In a large proportion of trials (42.5%) ‘DiD IT’ was unable to calculate a confidence interval around the central estimate. Furthermore, only 62.8% of the actual inject totals were within the 95% confidence interval for the parameter used as the central estimate for excess counts. Clearly, the 95% confidence interval should not be interpreted as representing very strong confidence that the actual number of excess cases is within the interval.

The main factor affecting the accuracy of ‘DiD IT’ estimates was the mean number of counts in the background data. Syndromic indicators which had higher daily counts, and therefore higher variance, resulted in bigger mean errors. The next most important factor affecting accuracy was whether an incident involved a single spike in cases or if the incident was spread over several days and/or districts. Incidents that were more spread out were less easy to quantify than single spikes in activity.

### What we already know

The difference-in-differences (DiD) method has been applied to answer many counterfactual questions in epidemiology [8, 9]. For instance, Branas at-al used DiD in a ten-year study to estimate the health benefits of greening vacant urban space [10], whilst Harper at-al used DiD to estimate the impact of marijuana laws [11].

Similarly, Xiongfeng et-al used DiD to estimate the impact on air pollution of a Smart Logistics policy [12]. In other environmental studies spatial difference-in-differences models have also been used to estimate impact whilst considering potential lags and spill-over across areas [13, 14]. Many DiD studies, even when daily data is collected are comparing differences between months or years of aggregated data, compared to the short periods for which ‘DiD IT’ is designed [15, 16]. Studies covering shorter periods enable rapid assessments of public health interventions, such as introducing and removing containment measures introduced following the SARS-CoV-2/Covid-19 pandemic [17, 18].

### Implications

The validation trials presented here have shown that ‘DiD IT’ usually provides a good close estimate to the number of excess cases in a specific location. Furthermore, where confidence intervals were calculated the majority were sufficiently narrow to provide useful information to decision-makers. However, further work is needed to provide intervals that are easier to interpret for users.

‘DiD IT’ has been tested across a range of syndromic systems and indicators utilised in UKHSA, and dates and different types of incidents represented by different synthetic injects. Thus, we can be confident that ‘DiD IT’ will work for a wide range of incidents. The main factor affecting the absolute size of errors was the mean number of daily counts in the background data. It is to be expected with count data that variance and therefore the absolute size of errors will increase with volume. We found that ‘DiD IT’ was less likely to provide an estimate for the GP in-hours system, which is probably due to the spareness of data at weekends and during public holidays.

‘DiD IT’ was less accurate when trying to quantify an incident spread over several days or locations but with a central peak in activity. However, the accuracy was still high when trying to estimate just the peak day or central district. Therefore, ‘DiD IT’ will be most useful when focussed on the centre of any known exposure, with accuracy likely to diminish if trying to estimate any additional spill over to neighbouring districts or days. Reassuringly, estimating the extra cases in the central peak does not seem to be adversely affected by failing to exclude spill over effects from controls.

We did not find that including a washout period or excluding neighbours to exposed districts improved the accuracy of ‘DiD IT’. In practice, including as many locations as possible in controls and using a full week for the control period gave better results than trying to replicate the size of exposure.

### Limitations

Syndromic surveillance was initially designed for national surveillance where higher aggregated data provides more confidence around emerging trends. When we drill down to local areas the data becomes sparse with many zero counts. By contrast, any significant incident may appear large and obvious within the data. Therefore, it is possible that the synthetic injects we are using provide too easy a target for ‘DiD IT’ and it is not surprising the accuracy is good. However, to include injects smaller than a peak of five cases in a day would mean we are simulating events that are in general too small to be relevant for public health surveillance. Furthermore, we are only considering incidents where a minimum number of cases present to health care.

We cannot estimate the number of people affected who do not seek care and thus do not appear in the syndromic data.

Interpretation of low or negative estimates from ‘DiD IT’ should not be taken as reassurance that an incident has had no impact unless we are confident that the syndromic systems have good coverage in the exposure location. Therefore, real-time assessments of system coverage need to accompany reports.

With some types of incidents it is quite likely that the exact timing of the exposure duration or its location are uncertain. Although, we have shown ‘DiD IT’ is robust to some exposure spread it will not be accurate if the peak is miss-identified. For example, the timing of patients presenting to health care may be affected by unknown factors causing a lag between exposure and presentation with symptoms.

‘DiD IT’ uses unaffected locations and dates as controls. An alternative method would be to use syndromic indicators that should be unrelated to the incident as negative controls, for instance gastrointestinal symptoms when considering air pollution [19].

‘DiD IT’ provides a single central estimate for the number of excess cases due to an incident. However, incident directors may want to know how an incident is evolving over time, or whether some locations are more adversely affected than others.

Furthermore spatial or temporal changes in an incident may not be linear or heterogenous across ages etc. Further work is needed before ‘DiD IT’ could provide this level of detailed analysis.

‘DiD IT’ focuses on a count of excess cases, however teams managing the incident response may find it more useful to see an estimate for changes in local incidence. Although calculating incidence rates is possible, care needs to be taken with syndromic data because many different diseases and illnesses present with similar symptoms.

### Future work

The application of ‘DiD IT’ alongside the existing use of syndromic surveillance for will further enhance the usability of syndromic data for supporting the response to incidents or emergencies. However, as highlighted in this validation, further work is needed to replace confidence intervals with data or prediction intervals [20] that provide users with meaningful information about the uncertainty of estimates. One approach would be to use our trial data to train an algorithm to construct intervals that capture a known percentage of inject totals. The utility of ‘DiD IT’ depends not just in providing accurate estimates but also in providing confidence that the estimates are accurate.

A further validation could include testing ‘DiD IT’ against real known incidents, particularly those where we know how many people presented to health care. In additional to further validating ‘DiD IT’, this approach could also evaluate the detection capabilities of syndromic surveillance systems. For example, if we could identify norovirus outbreaks in known locations and dates we could test whether the outbreaks resulted in a measurable increase in syndromic indicators. Furthermore, a meta-analysis could provide an overall estimate for the typical impact of local norovirus outbreaks.

## Acknowledgements

We acknowledge the UK Health Security Agency Real-time Syndromic Surveillance Team for technical expertise in delivering the daily syndromic service. We also thank syndromic data providers: NHS 111 and NHS Digital; TPP and participating TPP practices supporting GP in-hours; Advanced and participating GP OOH providers; Association of Ambulance Chief Executives and NHS Ambulance Trusts.

RAM and AJE receive support from the National Institute for Health Research (NIHR) Health Protection Research Unit (HPRU) in Emergency Preparedness and Response at Kings College London. AJE receives support from the NIHR HPRU in Gastrointestinal Infections at the University of Liverpool. The views expressed are those of the author(s) and not necessarily those of the NIHR, UK Health Security Agency or the Department of Health and Social Care.

## Ethical statement

All data used in this study were anonymised. The UKHSA has access to a range of data sources under Regulation 3 (Health Protection) of the Health Service (Control of Patient Information) Regulations 2002. This study therefore did not require specific ethics approval.

## Data Availability Statement

Applications for requests to access relevant anonymised data included in this study should be submitted to the UKHSA Office for Data Release. Available at: https://www.gov.uk/government/publications/accessing-ukhsa-protected-data.

## Notes

### Competing Interest Statement

The authors have declared no competing interest.

### Funding Statement

No specific funding was received - this work was carried out within the remit of UKHSA.

## References

1. Morbey R, et al. (2023) DiD IT: A differences-in-differences investigation tool to quantify the impact of local incidents on public health using real-time syndromic surveillance health data. Epidemiology & Infection, 1-11.

2. Yoon PW, Ising AI and Gunn JE (2017) Using Syndromic Surveillance for All-Hazards Public Health Surveillance: Successes, Challenges, and the Future. Public Health Reports 132, 3S–6S.

3. Agency UHS. Syndromic surveillance: systems and analyses. Available at https://www.gov.uk/government/collections/syndromic-surveillance-systems-and-analyses (Accessed 22/06/2023 2023).

4. Smith GE, et al. (2019) Syndromic surveillance: two decades experience of sustainable systems - its people not just data! Epidemiology and Infection 147, e101.

5. Packer S, et al. (2022) The Utility of Ambulance Dispatch Call Syndromic Surveillance for Detecting and Assessing the Health Impact of Extreme Weather Events in England. International Journal of Environmental Research and Public Health 19, 3876.

6. Harcourt SE, et al. (2016) Developing and validating a new national remote health advice syndromic surveillance system in England. Journal of Public Health 39, 184–192.

7. Harcourt SE, et al. (2012) Developing a new syndromic surveillance system for the London 2012 Olympic and Paralympic Games. Epidemiology and Infection 140, 2152–2156.

8. Tchetgen EP, C.; Richardon, D; (2023) Universal Difference-in-Differences for Causal Inference in Epidemiology. statME.

9. Strumpf Echskjs. Fixed Effects and Difference-in-differences. In: Oakes JMK, Jay S., ed. Methods in Social Epidemiology: John Wiley & Sons, 2017: pp. 341–368.

10. Branas CC, et al. (2011) A Difference-in-Differences Analysis of Health, Safety, and Greening Vacant Urban Space. American Journal of Epidemiology 174, 1296–1306.

11. Harper S, Strumpf EC and Kaufman JS (2012) Do Medical Marijuana Laws Increase Marijuana Use? Replication Study and Extension. Annals of Epidemiology 22, 207–212.

12. Pan X, et al. (2020) The effects of a Smart Logistics policy on carbon emissions in China: A difference-in-differences analysis. Transportation Research Part E: Logistics and Transportation Review 137, 101939.

13. Jia R, Shao S and Yang L (2021) High-speed rail and CO2 emissions in urban China: A spatial difference-in-differences approach. Energy Economics 99, 105271.

14. Zhao C, Wang K and Dong K (2023) How does innovative city policy break carbon lock-in? A spatial difference-in-differences analysis for China. Cities 136, 104249.

15. Renzi M, et al. (2019) Long-Term Exposure and Cause-Specific Mortality in the Latium Region (Italy): A Difference-in-Differences Approach. Environmental Health Perspectives 127, 067004.

16. Scapini VV, C.; Contreras, J.;. Social Crisis, Protests and Effects on Public Emergency Services: Econometric Analysis of the Chilean Social Outbreak. In: Passerini GG, F.; Lombardi, M., ed. Disaster Management and Human Health Risk VII: WIT Press, 2022.

17. Kosfeld R, et al. (2021) The Covid-19 containment effects of public health measures: A spatial difference-in-differences approach. Journal of Regional Science 61, 799–825.

18. Kapoor NR, et al. (2022) Effect of lifting COVID-19 restrictions on utilisation of primary care services in Nepal: a difference-in-differences analysis. British Medical Journal Open 12, e061849.

19. Morbey R, et al. (2022) Estimating the Impact of Air Pollution on Healthcare-Seeking Behaviour by Applying a Difference-in-Differences Method to Syndromic Surveillance Data. International Journal of Environmental Research and Public Health 19, 7097.

20. Barron EN and Del Greco JG. Confidence and Prediction Intervals. In: Barron EN, Del Greco JG, eds. Probability and Statistics for STEM: A Course in One Semester. Cham: Sprimger International Publishing, 2020: pp. 77–103.

